# Mutations in the U2 snRNA gene *RNU2-2P* cause a severe neurodevelopmental disorder with prominent epilepsy

**DOI:** 10.1101/2024.09.03.24312863

**Authors:** Daniel Greene, Koenraad De Wispelaere, Jon Lees, Andrea Katrinecz, Sonia Pascoal, Emma Hales, Marta Codina-Solà, Irene Valenzuela, Eduardo F. Tizzano, Giles Atton, Deirdre Donnelly, Nicola Foulds, Joanna Jarvis, Shane McKee, Michael O’Donoghue, Mohnish Suri, Pradeep Vasudevan, Kathy Stirrups, Natasha P. Morgan, Kathleen Freson, Andrew D. Mumford, Ernest Turro

## Abstract

The major spliceosome comprises the five snRNAs U1, U2, U4, U5 and U6. We recently showed that mutations in *RNU4-*2, which encodes U4 snRNA, cause one of the most prevalent monogenic neurodevelopmental disorders. Here, we report that recurrent germline mutations in *RNU2-2P*, a 191bp gene encoding U2 snRNA, are responsible for a related disorder. By genetic association, we implicated recurrent *de novo* single nucleotide mutations at nucleotide positions 4 and 35 of *RNU2-2P* among nine cases. We replicated this finding in six additional cases, bringing the total to 15. The disorder is characterized by intellectual disability, neurodevelopmental delay, autistic behavior, microcephaly, hypotonia, epilepsy and hyperventilation. All cases display a severe and complex seizure phenotype. Our findings cement the role of major spliceosomal snRNAs in the etiologies of neurodevelopmental disorders.

---

Over 1,400 genes have been established as etiological for neurodevelopmental disorders (NDDs) featuring intellectual disability, of which only 10 are non-coding^1^. Three of these ten non-coding genes—*RNU4ATAC*, *RNU12* and *RNU4-2*—encode small nuclear RNAs (snRNAs) that play crucial roles in orchestrating gene splicing. Variants in *RNU4ATAC* are responsible for microcephalic osteodysplastic primordial dwarfism, type I (MOPDI)^2^, Roifman syndrome^3^ and Lowry syndrome^4^, while variants in *RNU12* cause early onset cerebellar ataxia^5^. These pathologies are inherited in an autosomal recessive manner. Both *RNU4ATAC* and *RNU12* encode components of the minor spliceosome, a molecular complex that orchestrates splicing for about 0.35% of all splice junctions in humans^6^. However, more than 99% of introns are spliced by the major spliceosome. Recently, we reported that *de novo* mutations in *RNU4-2*, which encodes the U4 snRNA component of the major spliceosome, cause one of the most prevalent monogenic NDDs^7^. The discovery was later published independently by a separate group^8^.

To explore whether other non-coding genes might also be causal for NDDs, we performed a refined statistical analysis of the 100,000 Genomes Project (100KGP) data in the National Genomic Research Library (NGRL)^9^. Following a previously described approach^7,10^, we applied the BeviMed genetic association method^11^ to compare rare variant genotypes in 41,132 non-coding genes between 7,452 unrelated, unexplained cases annotated as having a neurodevelopmental abnormality (NDA) and 43,727 unrelated participants without an NDA. Importantly, while our previous analyses filtered out SNVs having a combined annotation-dependent depletion (CADD) score <10, our present analysis relaxed this threshold as, in non-coding genes, it has been shown that CADD scores are a poor discriminant of variant pathogenicity^12^.

Our analysis yielded only two genes with a posterior probability of association (PPA) with NDA >0.5. *RNU4-2*, which encodes the snRNA U4, had a PPA≈1^7^, and *RNU2-2P*, which encodes the snRNA U2, had a PPA=0.97. The novel association with *RNU2-2P* depended on inclusion of variants with CADD scores ≤ 10 (**Extended Data Fig. 1**). Conditional on the association, two variants, at nucleotide positions 4 and 35, had a BeviMed posterior probability of pathogenicity (PPP) >0.5 (**Fig. 1a**). The 9 NDA cases with either of the variants had a significantly greater phenotypic homogeneity based on Human Phenotype Ontology (HPO) terms than expected under random selection of 9 NDA cases from unexplained and unrelated NDA study participants in the 100KGP (**Fig. 1b**), supporting causality for a distinct NDD. Although *RNU2-2P* is annotated as a pseudogene in bioinformatics databases^13^, it is the most highly expressed of the 71 known homologs of U2 across tissues, including brain and non-brain tissues, blood cells (**Fig. 1c**, **Extended Data Fig. 2**) and cell lines^14^. Moreover, *RNU2-2P* resides in a 5’ untranslated exon of *WDR74* that had previously been identified as being enriched for hotspot mutations in cancer, although the existence of *RNU2-2P* at that locus was not known at the time^15^. More recently, it has been shown that *RNU2-2P* is a functional gene that is transcribed independently of *WDR74* and carries recurrent somatic mutations (n.28C>T and n.28C>G) that drive B-cell derived tumors, prostate cancers and pancreatic cancers^16^.

**Fig. 1.**
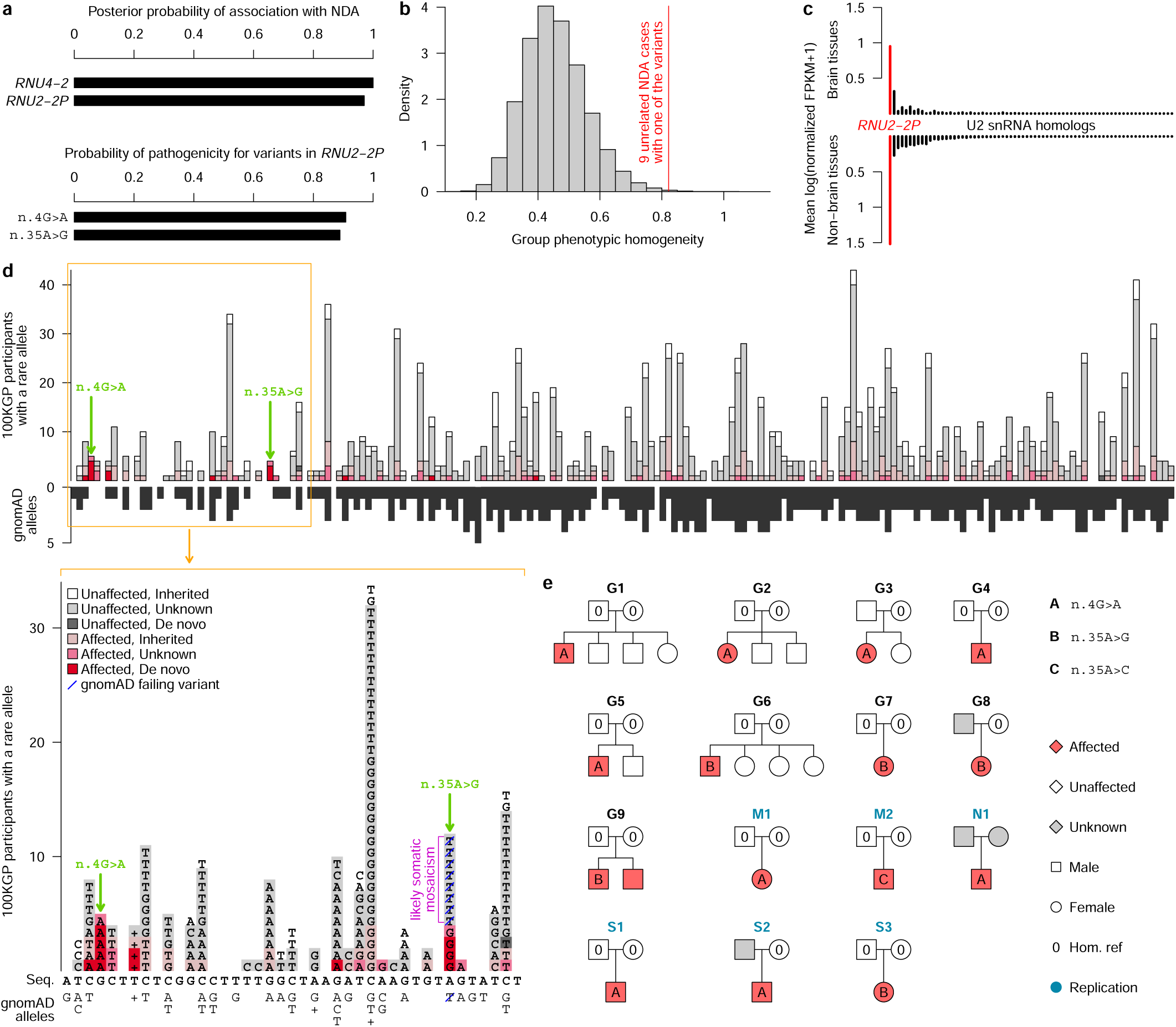
Discovery and replication of *RNU2-2P* as an etiological gene for a novel NDD. **a**, BeviMed PPAs between each of *RNU4-2* and *RNU2-2P* and NDA. All other non-coding genes had a PPA <0.5. Only two *RNU2-2P* variants had a conditional PPP >0.5: n.4G>A and n.35A>G. **b**, Distribution of phenotypic homogeneity scores (see Methods) for randomly selected sets of nine participants chosen from the 9,112 unrelated NDA cases. The actual score for the nine ID cases with one of the two *RNU2-2P* variants having a PPP >0.5 is indicated with a red line. **c**, Bar plot of the mean (over brain and non-brain tissues in GTEx) of the mean (over samples) log expression of each of the 71 annotated U2 snRNA gene homologs. The bars are ordered by the sum over tissue categories. The bar corresponding to *RNU2-2P* is highlighted in red, showing it is by far the most highly expressed homolog. **d**, Above, for each of the 191 bases of the *RNU2-2P* gene, the number of participants with a rare allele at that position, stratified by affection status and inheritance information of the carried rare allele. The bases corresponding to the two variants with a PPP>0.5 are indicated with green arrows. The black bars indicate the number of distinct alternate alleles in gnomAD at each position (multiple insertions and multiple deletions at a given position each count as one). Underneath, the data corresponding to nucleotide positions 1 to 41 are shown in greater detail: above and below the reverse complement of the *RNU2-2P* sequence (Seq.), the alternate alleles in 100KGP participants and the distinct alleles in gnomAD are shown, respectively, where ‘+’ indicates insertions and diagonal blue dash indicates variants that fail QC in gnomAD. **e**, Pedigrees for participants with a rare alternate allele n.4 or n.35 of *RNU2-2P*. Pedigrees used for discovery have a “G” prefix and are labeled in black. Pedigrees used for replication in the 100KGP, in the NBR or in the GMS have an “M”, “N” or “S” prefix, respectively, and are labeled in blue.

The two germline variants with a high PPP, n.4G>A and n.35A>G, are located in a genomic locus spanning a region of approximately 40 nucleotides at the 5’ end of the 191bp *RNU2-2P* gene. The locus has a markedly reduced density of variation in gnomAD^17^, which is consistent with the effects of negative selection (**Fig. 1d**). Published secondary structure data of the U2 snRNA show that n.4 is a crucial interactor of U6 within the pre-B and B complexes, while n.35 binds the branch sites of introns^18^ (**Extended Data Fig. 3**). Trio sequencing of four of the five cases with n.4G>A and three of the four cases with n.35A>G showed the variants were *de novo* in each case. A variant with a different alternate allele at nucleotide 35, n.35A>T, was called in eight unaffected participants but failed quality control (QC) in gnomAD (**Fig. 1d**). Analysis of whole-genome sequencing (WGS) allele-specific coverage and Sanger sequencing data suggested that n.35A>G is a germline variant, but that n.35A>T is a recurring somatic mosaic variant. This somatic variant is observed only in individuals over the age of 40, which is consistent with clonal hematopoiesis (**Extended Data Fig. 4**).

To replicate our findings, we examined three additional rare disease collections: a component of the 100KGP not included in the discovery dataset (10,373 participants, of which 1,736 have an NDA), the NIHR BioResource-Rare Diseases (NBR)^19^ (7,388 participants, of which 731 have an NDA), and the UK’s Genomic Medicine Service (GMS) data (32,030 participants, of which 6,469 have an NDA). We identified a further six cases in these replication collections with *de novo* variants and no unaffected carriers of either variant. Four cases had n.4G>A, one case had n.35A>G and one case had a different alternate allele at nucleotide 35: n.35A>C. Although this case was the only individual harboring n.35A>C, modeling of the interactions between U2 snRNA and canonical branch site sequences suggested that n.35A>C has a destabilizing effect on binding that is greater than that of the n.35A>G variant observed in five cases and in many cases similar in magnitude to that of the n.4G>A variant with respect to its cognate partner U6 (**Extended Data Fig. 5**). All of these variants were called confidently by WGS (**Extended Data Fig. 6**). In the 100KGP, the prevalence of cases with these *RNU2-2P* mutations was five times smaller than that of *RNU4-2* syndrome, yet still more prevalent than all but 29 of the ~1,400 known etiological genes for intellectual disability (**Extended Data Fig. 7**).

Analysis of the HPO terms assigned to the nine uniformly phenotyped 100KGP cases revealed that 100% had ‘Intellectual disability’ and ‘Global developmental delay’, 89% had ‘Delayed speech and language development’, 78% had ‘Motor delay’ and 56% had ‘Autistic behavior’, in line with frequencies in NDA cases generally (**Fig. 2**). However, certain terms were enriched in *RNU2-2P* cases: ‘Seizure’ was present in 89% of cases (vs. 27% in other NDA cases, Bonferroni-adjusted (BA) *P*=2.44×10^-3^), ‘Microcephaly’ in 78% of cases (vs. 18%, BA *P*=1.62×10^-3^), ‘Generalized hypotonia’ in 56% of cases (vs. 13%, BA *P*=3.56×10^-2^), ‘Severe global developmental delay’ in 44% (vs. 2.7%, BA *P*=8.89×10^-4^) and ‘Hyperventilation’ in 33% of cases (vs. 0.16%, BA *P*=7.56×10^-6^). No HPO terms were significantly underrepresented in the *RNU2-2P* cases. Of the terms that were enriched amongst cases with *RNU4-2* syndrome, ‘Seizure’, ‘Microcephaly’, ‘Generalized hypotonia’ were also enriched in *RNU2-2P* cases.

**Fig. 2.**
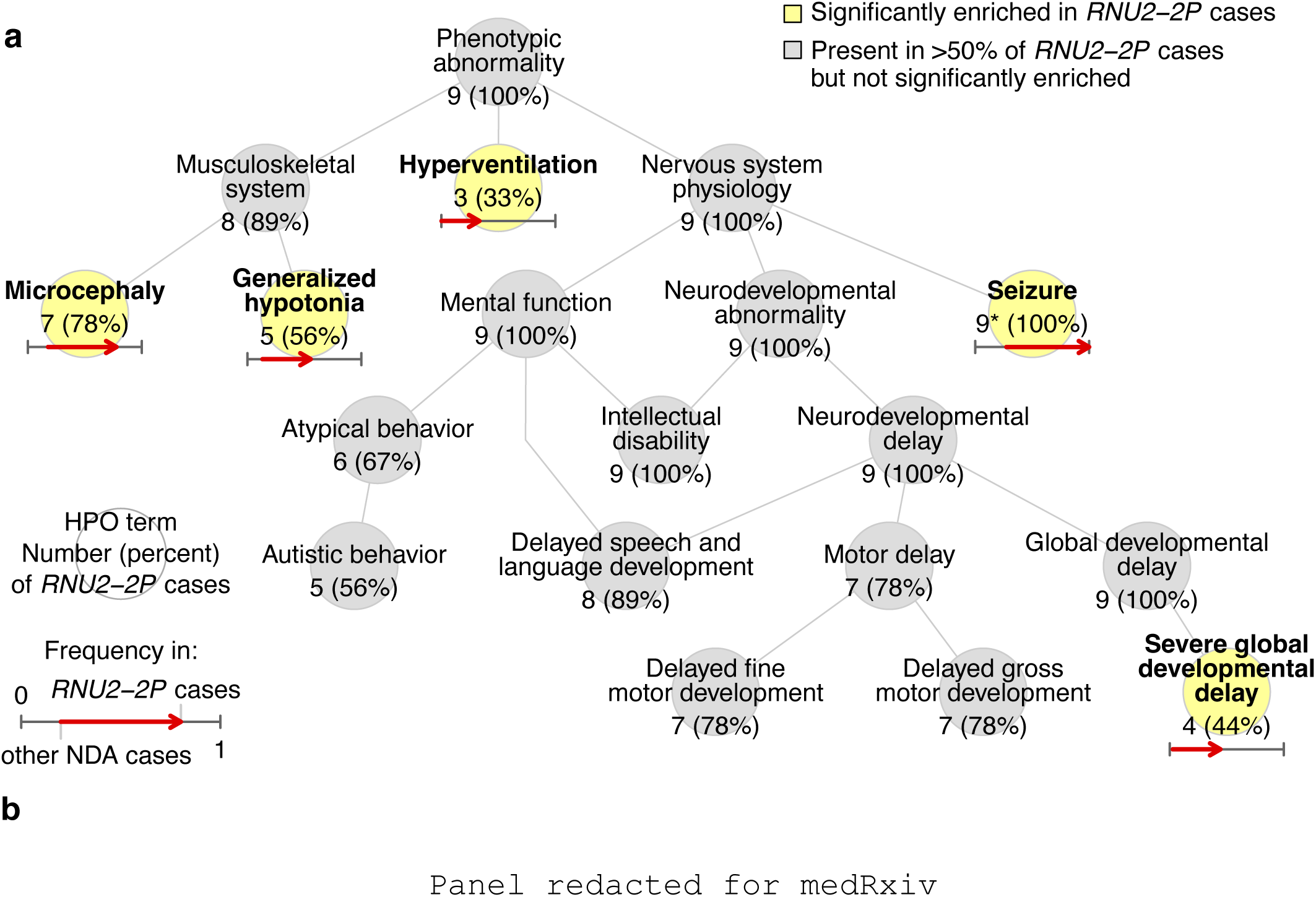
Phenotypic and mechanistic characterization. **a**, Graph showing the ‘is-a’ relationships among HPO terms present in at least three of the nine NDA-coded *RNU2-2P* cases in the discovery collection or significantly enriched among them relative to 9,112 unrelated NDA-coded participants of the 100KGP. The significantly overrepresented terms are highlighted. For each term, the number of cases with the term and the percentage that number represents out of nine is shown. For each overrepresented term, the proportion of NDA-coded participants with the term and the proportion of NDA-coded *RNU2-2P* cases with the term are represented as the horizontal coordinate of the base and the head of an arrow, respectively. * Only eight of the nine (89%) of the cases had the ‘Seizure’ HPO term in the NGRL. However, epilepsy was confirmed in the case without the HPO term by inspecting the case’s electronic health record. **b**, Clinical photographs of the cases from pedigrees G1, G4 and S3. The cases share the common features of long palpebral fissures with slight eversion of the lateral lower lids, long eyelashes, broad nasal root, large low set ears, wide mouth and wide spaced teeth. The approximate ages of the cases when the photographs were taken are shown.

However, ‘Severe global developmental delay’ and ‘Hyperventilation’ were only enriched in *RNU2-2P* cases, suggesting that these may be differentiating phenotypic features. Strikingly, three *RNU2-2P* cases were coded with the seldom used ‘Hyperventilation’ term by three independent clinicians.

Detailed clinical vignettes for the cases in pedigrees G1, G2, G4, M2 and S3 are provided in **Supplementary Information**. These indicate that the neurodevelopmental phenotype caused by the *RNU2-2P* variants typically manifests from three to six months of age but is progressive, frequently severe and is accompanied by characteristic dysmorphic features (**Fig. 2b**). All the cases displayed prominent epilepsy, usually from the first few months of life, and seizures were severe and pharmaco-resistant. Seizures were characteristically complex and included spasms, tonic, tonic clonic, myoclonic and absence types, classified in some probands as Lennox-Gastaut syndrome. These features distinguish the *RNU2-2P* cases from previously reported cases with *RNU4-2* syndrome, in which the developmental phenotype is reported as less severe, the dysmorphic features different and epilepsy typically later in onset, less severe and more commonly focal^7,8,20^. Extraordinarily, case M2 also harbored a *de novo* truncating variant in *SPEN* predicted to cause Radio-Tartalgia syndrome.^21^ However, the case had short stature (<0.4th centile) and microcephaly (≪0.4th centile), which are not characteristic of Radio-Tartalgia syndrome, whilst also having a morphology that more closely resembled other *RNU2-2P* patients than Radio-Tartalgia patients.

Analysis of uniquely aligned reads at heterozygous sites in whole blood RNA-seq data revealed that both alleles of *RNU2-2P* are expressed robustly in cases (**Extended Data Fig. 8**). However, gene expression analysis comparing the five cases to 495 unrelated unexplained participants with an NDA did not reveal numbers of differentially expressed genes or differentially used splice junctions above what was expected under random permutation of case labels, suggesting that transcriptomic analysis of other tissue types will be required to uncover the underlying molecular mediator of disease.

Mutant *RNU2-2P* mice do not exist but, interestingly, mice with a homozygous 5bp deletion in the U2 ortholog *Rnu2-8* present with ataxia and neurodegeneration^22^. Transcriptomic analysis of the mutant cerebellum detected alternative splicing, mainly increased retention of short introns. While it remains unclear how this splicing defect causes neuron death, it has been hypothesized that several retained introns contain premature translation termination codons that might trigger the nonsense-mediated mRNA-decay (NMD) pathway. Interestingly, we and others have shown that the human disorders caused by recessive inheritance of variants in *RNU4ATAC* and *RNU12* result in minor intron retention in blood cells and fibroblasts^2,3,5,23^. In contrast, we have been unable to detect a significant and reproducible large-scale intron retention defect in either blood cells or fibroblasts of patients with dominant germline variants in the major spliceosome genes *RNU4-2* and *RNU2-2P.* Although a recent study described systematic disruption of 5’ splice site usage in whole blood of some patients with *de novo RNU4-2* variants^8^, RNA-seq of fibroblasts in a separate case study could not detect any defect in splicing^20^. Moreover, transcriptomic analysis of primary hematological tumors and cell lines infected with vectors expressing the U2 c.28C>T *RNU2-2P* mutation did not reveal any significant differences in splicing^16^. Therefore, further studies are required to understand how *RNU4-2* and *RNU2-2P* mutations affect splicing. It might be that, in contrast to recessive disorders, it is challenging to detect splicing defects in these newly discovered dominant disorders because wild type transcripts are expressed in combination with misspliced transcripts from the same gene that are subjected to NMD. In certain cell types, the effects of NMD might be overcome such that the overall expression levels of mRNAs remain unchanged, due to rapid mRNA turnover and dosage compensation^24^. However, certain cell types, such as stem cells, which we have not yet been able to study, might be more sensitive to high NMD dosage than differentiated cells. Neuronal stem cell and mouse models of *RNU4-2* and *RNU2-2P* pathologies will be needed to resolve these mechanistic questions.

## Acknowledgements

This research was made possible through access to data in the National Genomic Research Library, which is managed by Genomics England Limited (a wholly owned company of the Department of Health and Social Care). The National Genomic Research Library holds data provided by patients and collected by the NHS as part of their care and data collected as part of their participation in research. The National Genomic Research Library is funded by the National Institute for Health Research and NHS England. The Wellcome Trust, Cancer Research UK and the Medical Research Council have also funded research infrastructure. We thank NIHR BioResource volunteers for their participation, and gratefully acknowledge NIHR BioResource centers, NHS Trusts and staff for their contribution. We thank the National Institute for Health and Care Research, NHS Blood and Transplant, and Health Data Research UK as part of the Digital Innovation Hub Programme. The views expressed are those of the author(s) and not necessarily those of the NHS, the NIHR or the Department of Health and Social Care. K.F. was supported by Katholieke Universiteit (KU) Leuven Special Research Fund (BOF) (C14/19/096 and C14/23/121), Research Foundation – Flanders (G072921N) and NIH award R01HL161365. K.D.W. was supported by the Belgian American Education Foundation and NIH award R01HL161365. A.M. was supported by NIH award R01HL161365. D.G., and E.T. were supported by NIH awards R01HL161365 and R03HD111492 and E.T. was further supported by the Lowy Foundation USA.

## Author contributions

D.G. conducted statistical and bioinformatic analyses and cowrote the paper. K. D. W. analyzed RNA-seq data, generated expression heatmaps and made the illustration showing molecular interactions. J.L. modeled free energies of association. A.K. processed NBR RNA-seq data. S.P. performed PCR and Sanger sequencing. E.H. oversaw recruitment to the NBR RNA-seq project. M.C-S., I.V. and E.F.T. designed primers, selected cases for sequencing and provided early access to detailed phenotype data on *RNU4-2* cases for comparative analysis. G.A., D.D., N.F., J.J., S.McK., M.O’D., M.S., and P.V. obtained consent and provided detailed phenotype information. M.O’D. also provided expert clinical interpretation. K.S. and N.M. oversaw the NBR RNA-seq study. K.F. provided biological interpretation and cowrote the paper. A.M. coordinated clinical contacts, provided clinical and biological interpretation and cowrote the paper. E.T. oversaw the study and cowrote the paper.

## Methods

### Enrollment

The enrollment criteria for participants in the NGRL are available from the Genomics England website^25^. The enrollment criteria for NBR participants are given in ref.^19^.

### Genetic association analysis

The genetic association analysis was conducted as described previously^7,10^, except that variants were not thresholded on CADD score. Cases comprised all of the 9,112 unrelated cases in the 100KGP included in the merged variant call format (VCF) file provided by the 100KGP who were annotated with the NDA HPO, while the controls comprised all of the 40,937 unrelated participants in the merged VCF who were not assigned the NDA term.

### Phenotypic homogeneity analysis

To assess the phenotypic homogeneity of the nine participants in the discovery collection with n.4G>A or n.35A>G in *RNU2-2P*, we computed a phenotype homogeneity score for that group with respect to unexplained and unrelated NDA study participants. We calculated this score using the ‘get_sim_grid’ and ‘get_sim_p’ functions from the ontologySimilarity R package,^26^ as previously described^7^. We then obtained a Monte Carlo *P*-value as the proportion of random sets of nine unexplained unrelated NDA cases having a homogeneity that was greater than or equal to the homogeneity score of the group carrying either of the *RNU2-2P* variants.

### Analysis of HPO terms

To identify enriched or depleted HPO terms among the nine NDA-annotated cases with n.4G>A or n.35A>G in *RNU2-2P* in the discovery collection, compared with unrelated NDA-coded participants without either of these two variants, we computed *P*-values of association using Fisher’s two-sided exact test. We only tested enrichment for terms attached to at least three of the nine cases and which belonged to the set of non-redundant terms at each level of frequency among the cases. To account for multiple comparisons, we adjusted the *P*-values by multiplying them by the number of tests. An adjusted *P*-value <0.05 was deemed statistically significant. To visualize both common and distinctive HPO terms for *RNU2-2P* cases, we selected terms that were either statistically significant or present in at least 50% of the cases, removed redundant terms at each level of frequency among the nine cases, and arranged the terms along with a non-redundant set of ancestral terms as a directed acyclic graph of is-a relations. These analyses were conducted using the ontologyX R packages^26^.

### Analysis of expression levels of U2 snRNA homologs

We downloaded transcript-level expression estimates from the GTEx portal, which were generated by genome alignment with STAR v2.5.3a and quantification with RSEM v1.3.0, accounting for alignments of reads to multiple regions of the reference genome. We summed the transcript-level expression estimates within genes by summing over isoforms. We then normalized the gene expression estimates using the TMM method and transformed them to FPKM values. Finally, we compute the mean log(FPKM + 1) within each tissue type. The NIHR BioResource – Rare Disease RNA Phenotyping Study is a multicentre multiomics study of approximately 1,000 patients. It consists of RNA sequencing and proteomics of platelets, neutrophils, monocytes and CD4+ T-cells with WGS for each participant where possible. We aligned the NIHR BioResource blood cell RNA-seq data to the genome using STAR v2.7.10a and quantified expression estimates with RSEM v1.3.1. The gene level expression estimates were computed as described above for each blood cell type.

### Mosaicism analysis

To compute the proportions of WGS reads supporting alternate alleles, we extracted the sequencing depth and the number of reads supporting each alternate allele at n.4 and n.35 of *RNU2-2P* from BAM files using ‘samtools mpileup’ (samtools version 1.16.1) with default settings.

### Sanger sequencing

We used the following primers to amplify genomic DNA containing the *RNU2-2P* gene prior to Sanger sequencing: forward primer: CCAATCCCAGGATCCTAAAAA; reverse primer: GAAGACCACATGGAGATACTACG. The amplified fragments correspond to chr11:62841419– 62842071 in version GRCh38 of the human reference genome.

### Modeling free energies of association

We calculated the free energy of duplex formation ΔG^27^ of duplex formation with U6 and with branch site sequences for wildtype and mutant U2 using the RNA.fold_compound.eval_structure function in the ViennaRNA (v2.6.4) python package. This allowed us to calculate the difference in stability change on mutation, ΔΔG. For U2-2P, we already know from the literature the U2 RNA duplex pairings formed with the branch point and U6 sequences (allowing us to pre-specify the structure and evaluate ΔG from this).

### RNA-seq data analysis

We performed QC on RNA-seq data derived from the whole blood of 5,546 participants in the NGRL as follows. Based on visual inspection of QC parameter distributions, we filtered out samples with a percentage of RNA fragments larger than 200 bases (as measured by an Agilent Tapestation 4200) ≤ 65%, a total read count outside the range (108M, 592M), a genome mapping rate <0.85 or a high-quality read rate <0.9 (where reads were deemed to be of high quality if they aligned as proper pairs, had fewer than seven mismatches and had a mapping quality ≥ 60). After QC filtering, 5,165 samples remained for analysis, including the five cases with implicated variants in *RNU2-2P*. We assessed allele-specific expression in cases by counting genome-aligned RNA-seq reads overlapping heterozygous sites using ‘samtools mpileup’ (samtools version 1.16.1) with default settings. We selected 500 samples for differential gene expression and splice junction usage analysis by taking the five cases and 495 samples selected at random from those passing the QC criteria, and which belonged to unrelated NDA-coded individuals presently unexplained. We used DESeq2^28^ (version 1.44) to conduct both differential gene expression analysis and differential splice junction usage analysis. For differential gene expression analysis, we took the transcript quantifications generated by the Salmon software^29^ and aggregated them by gene using the ‘tximport’ BioConductor package^30^. For differential splice junction usage analysis we used the read counts of spliced reads supporting the junctions of the introns of the canonical transcripts of genes in Ensembl version 104. We only analyzed junctions that were observed (i.e., supported by at least one spliced read) in at least 99% of the 500 samples.

### Ethics

Participants of the 100KGP, the 100KGP Pilot Project and the GMS were enrolled to the National Genomic Research Library under a protocol approved by the East of England– Cambridge Central Research Ethics Committee (ref: 20/EE/0035). We obtained written informed consent to publish additional clinical data from a subset of the affected cases in the NGRL following local best practices. NBR participants were enrolled under a protocol approved by the East of England Cambridge South Research Ethics Committee (ref. 13/EE/0325).

## Data availability

Genetic and phenotypic data for the 100KGP study participants, the 100KGP Pilot study participants and the GMS participants are available through the Genomics England Research Environment via the application at https://www.genomicsengland.co.uk/join-a-gecip-domain. Data pertaining to: WGS data were obtained for 78,132 100KGP participants, 4,054 100KGP Pilot participants, 32,030 GMS participants (v3) and 13,037 NBR participants; HPO phenotype data from the ‘rare_diseases_participant_phenotype’ table (Main Programme v14), ‘observation’ table (GMS v3) and ‘hpo’ table (Rare Diseases Pilot v3); Specific Disease class data from the ‘rare_diseases_participant_disease’ table (Main Programme v13); ICD10 codes from the ‘hes_apc’ table (Main Programme v13); pedigree information from the ‘rare_diseases_pedigree_member’ table (Main Programme v13), ‘referral_participant’ table (GMS v3), and ‘pedigree’ table (Rare Diseases Pilot v3); explained/unexplained status of cases from the ‘gmc_exit_questionnaire’ tables (Main Programme v18, GMS v3). Accession codes for NBR data are given in ref.^19^. CADD v.1.5 (https://cadd.gs.washington.edu/), gnomAD v.3.0 (https://gnomad.broadinstitute.org/) and Ensembl v.104 (http://may2021.archive.ensembl.org/index.html) were used for variant annotation. Expression data for U2 snRNA homologs were extracted from the file ‘GTEx_Analysis_2017-06-05_v8_RSEMv1.3.0_transcript_expected_count.gct.gz’ available from the GTEx Portal. Data presented in this paper were requested from the Genomics England Airlock on August 13, 2024 at 3:39am British Summer Time (BST). The manuscript was submitted to the Genomics England Publication Committee on August 21, 2024 at 23:51 BST and approved for submission on August 27, 2024 at 15:52 BST.

## Code availability

Software packages rsvr 1.0, bcftools 1.16, samtools 1.9 and perl 5 were used to build the non-coding 100KGP Rareservoir. The Rareservoir software is available from https://github.com/turrogroup/rsvr. All R packages listed in the manuscript are available via the Comprehensive R Archive Network site (https://cran.r-project.org/) or Bioconductor (https://bioconductor.org). The ViennaRNA 2.6.4 package is available from the Python Package Index (https://pypi.org).

---

**Extended Data Fig. 1.**
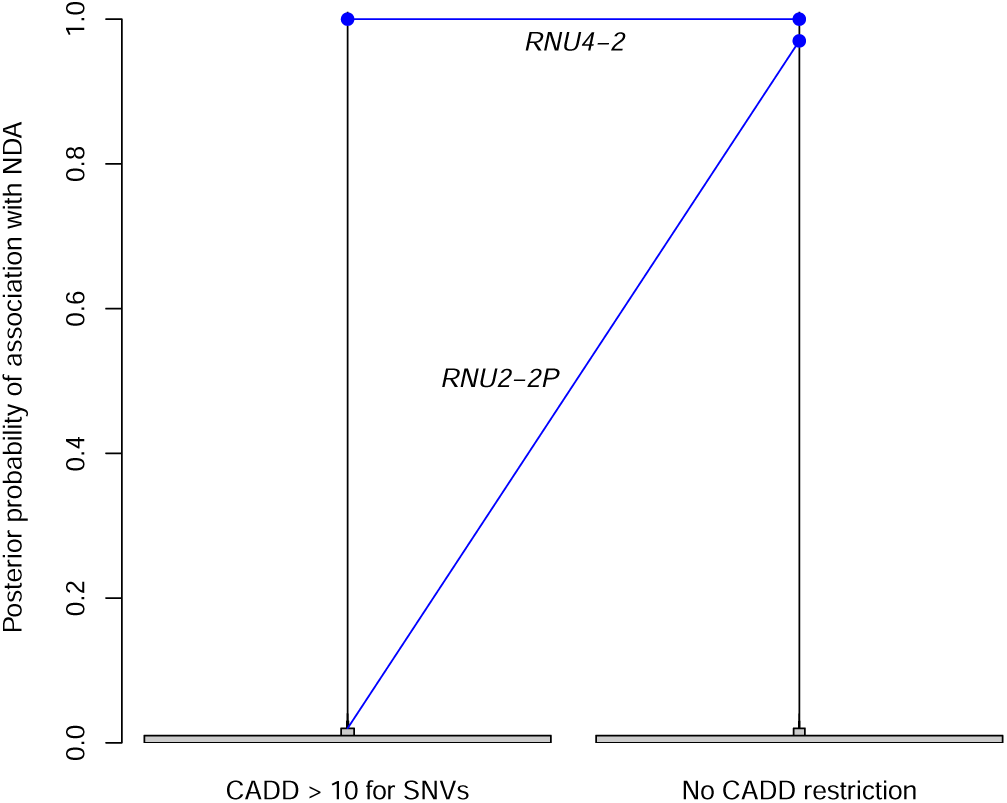
Effect on PPAs of relaxing the CADD score threshold. PPAs between non-coding genes and NDA, with and without filtering out variants with a CADD score <10.

**Extended Data Fig. 2.**
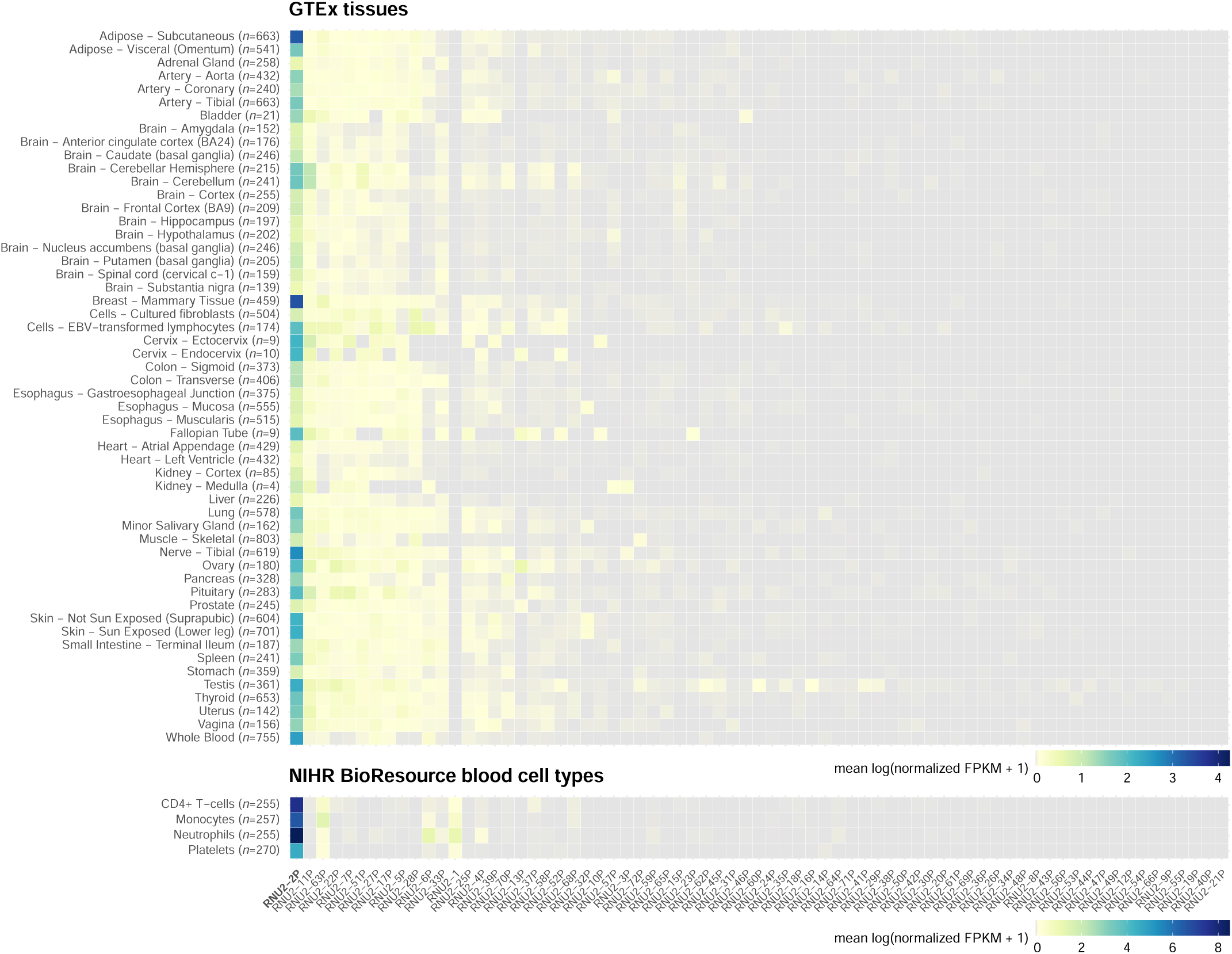
Expression heatmaps of U2 homologs. Mean log expression levels of the 71 U2 homologs in adult tissues sampled by GTEx (top) and four blood cell types sampled by the NIHR BioResource – Rare Disease RNA Phenotyping Study (bottom). The dynamic ranges of the two datasets differ because GTEx used a poly-A selection protocol^31^, which suppresses expression estimates of almost all non-coding genes, while the NIHR BioResource used a ribosomal RNA depletion protocol, which only suppresses the expression estimates of ribosomal RNA genes.

**Extended Data Fig. 3.**
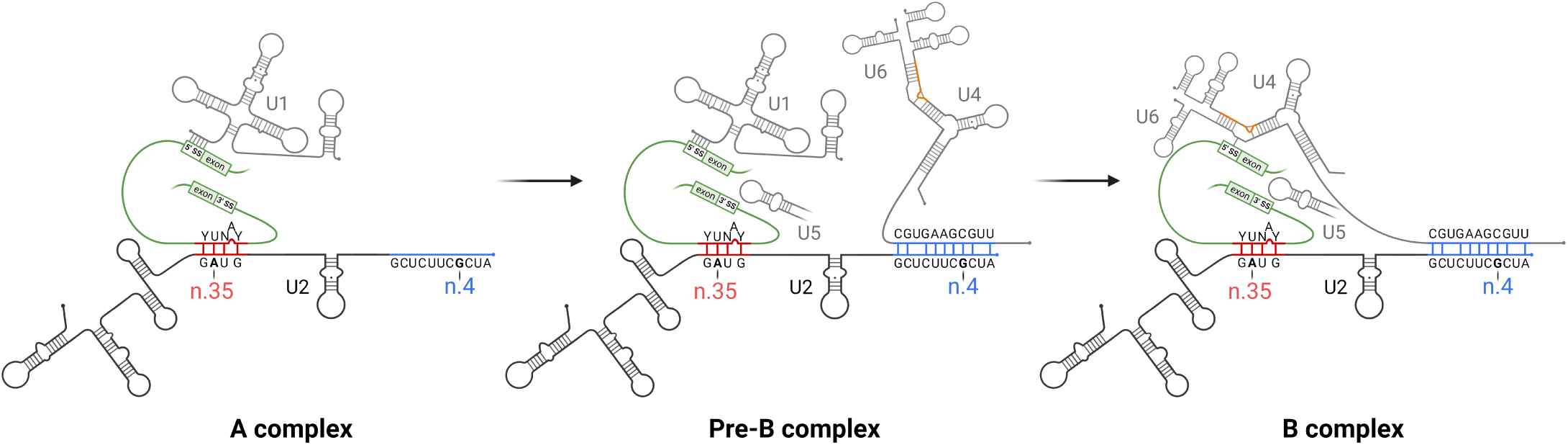
Location of the pathogenic variants in U2 snRNA within the major spliceosome. Assembly of the spliceosome A complex is initiated by binding of the intronic 5’ splice site (5’SS) to the U1 snRNA and the intronic branch site sequence to the U2 snRNA through Watson-Crick pairing of cognate ribonucleotides. The branch site sequence is depicted as the human YUNAY consensus motif (Y=C or T; N=any ribonucleotide), which interacts with the GUAG sequence at positions 33 to 36 in the U2 snRNA (depicted in red)^18^. The spliceosome pre-B complex is formed by incorporation of the U4/U6.U5 tri-small nuclear ribonucleoprotein (snRNP) complex that contains the U4, U5 and U6 snRNAs. This requires interactions between U5 snRNA and the 5’ and 3’ exons^32^ and further interactions between nucleotides near the 3’ end of the U6 snRNA and a cognate CGCUUCUCG sequence (nucleotides 3–11) close to the 5’ end of the U2 snRNA (depicted in blue)^33^. Tethering of U4/U6.U5 tri-snRNP to U2 within the spliceosome pre-B complex enables displacement of U1 to enable a new interaction between U6 snRNA with the 5’SS and reconfiguration of U4/U6.U5 tri-snRNP to form the catalytically active spliceosome B complex, which is a prerequisite for the splicing reaction^34^. The critical U6 snRNA region that interacts with the intronic 5’SS^35^ is maintained in correct orientation by conserved regions in the adjacent U4 snRNA (depicted in orange) which are the sites of destabilizing variants responsible for the recently described *RNU4-2* syndrome^7^. The variants responsible for *RNU2-2P* syndrome occur at critical interaction sites between U2 snRNA near n.4 and U6 snRNA and between U2 snRNA near n.35 and intronic branch sites. These interactions are necessary for intron recognition and the correct assembly of the catalytically active spliceosome B complex^36^.

**Extended Data Fig. 4.**
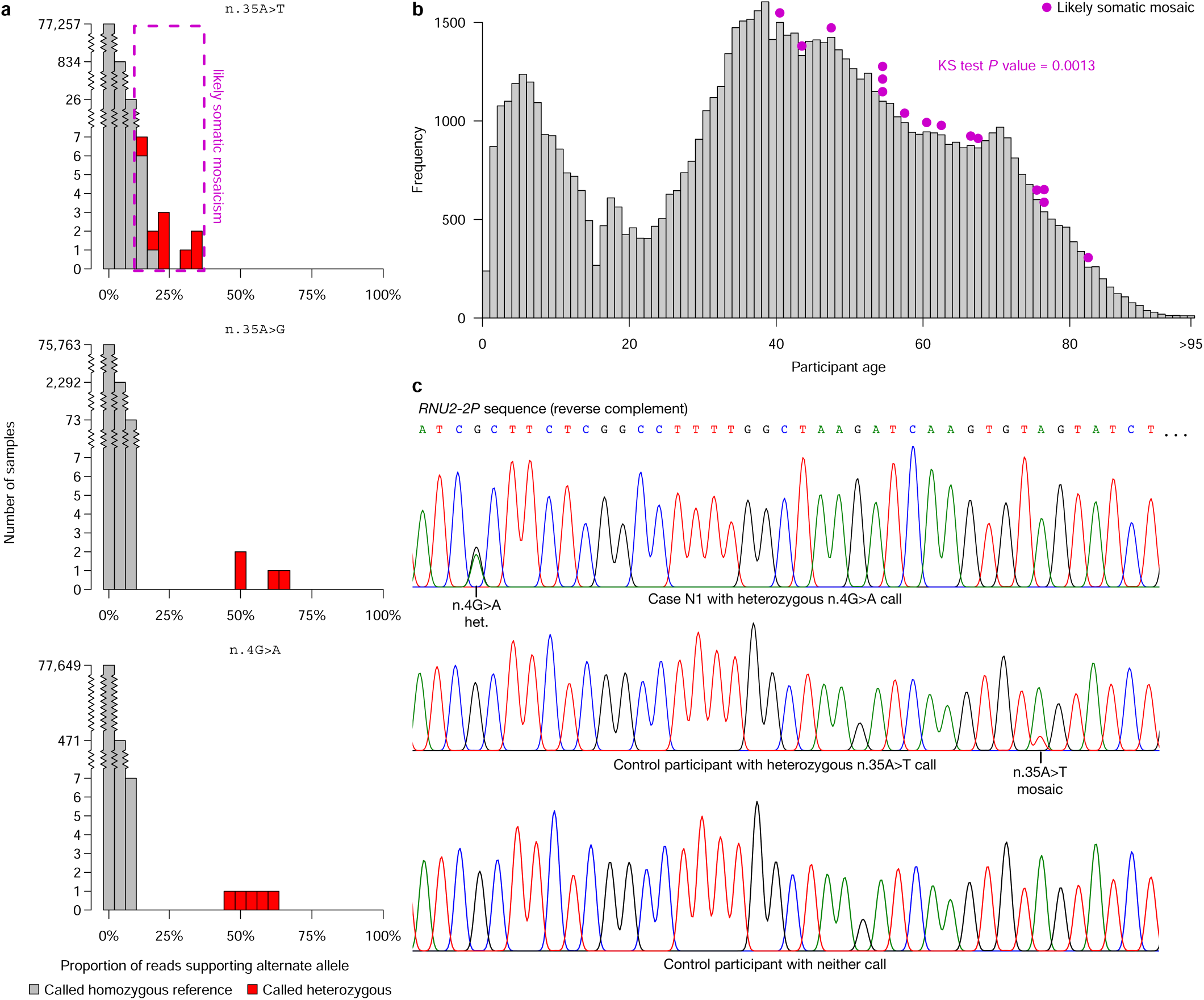
Mosaicism analysis. **a**, For each of the three rare variants at positions n.4 and n.35 of *RNU2-2P* called in the discovery collection, truncated bar charts showing the distribution of the proportions of reads supporting the alternate allele over participants, partitioned into 0% and all left-open intervals of size 4% up to 100%. In contrast to n.4G>A and n.35A>G, the reads in the eight participants with the n.35A>T heterozygous call exhibit a strong skew in favor of the reference allele. Furthermore, seven participants with a homozygous reference call at n.35 have at least 8% of aligned reads at that position supporting the ‘T’ allele, suggesting that n.35A>T is not a germline variant, but rather a low-frequency somatic mosaic variant. **b**, Histogram of age at enrollment of participants in the discovery collection. The purple points show the age at enrollment of study participants with at least 8% of aligned reads supporting the ‘T’ allele at n.35. These participants are significantly older than expected by chance (*P*=1.3×10^-3^, Kolmogorov-Smirnoff test). To comply with Genomics England’s rules on identifiability, all ages of at least 95 years are included in the same *x*=95 bin. **c**, Sanger sequencing traces from an NDA case (in pedigree N1) with the n.4G>A call, an unaffected participant with the n.35A>T call and a control with neither call, showing that n.4G>A is a germline variant while n.35A>T is a likely somatic mosaic variant.

**Extended Data Fig. 5.**
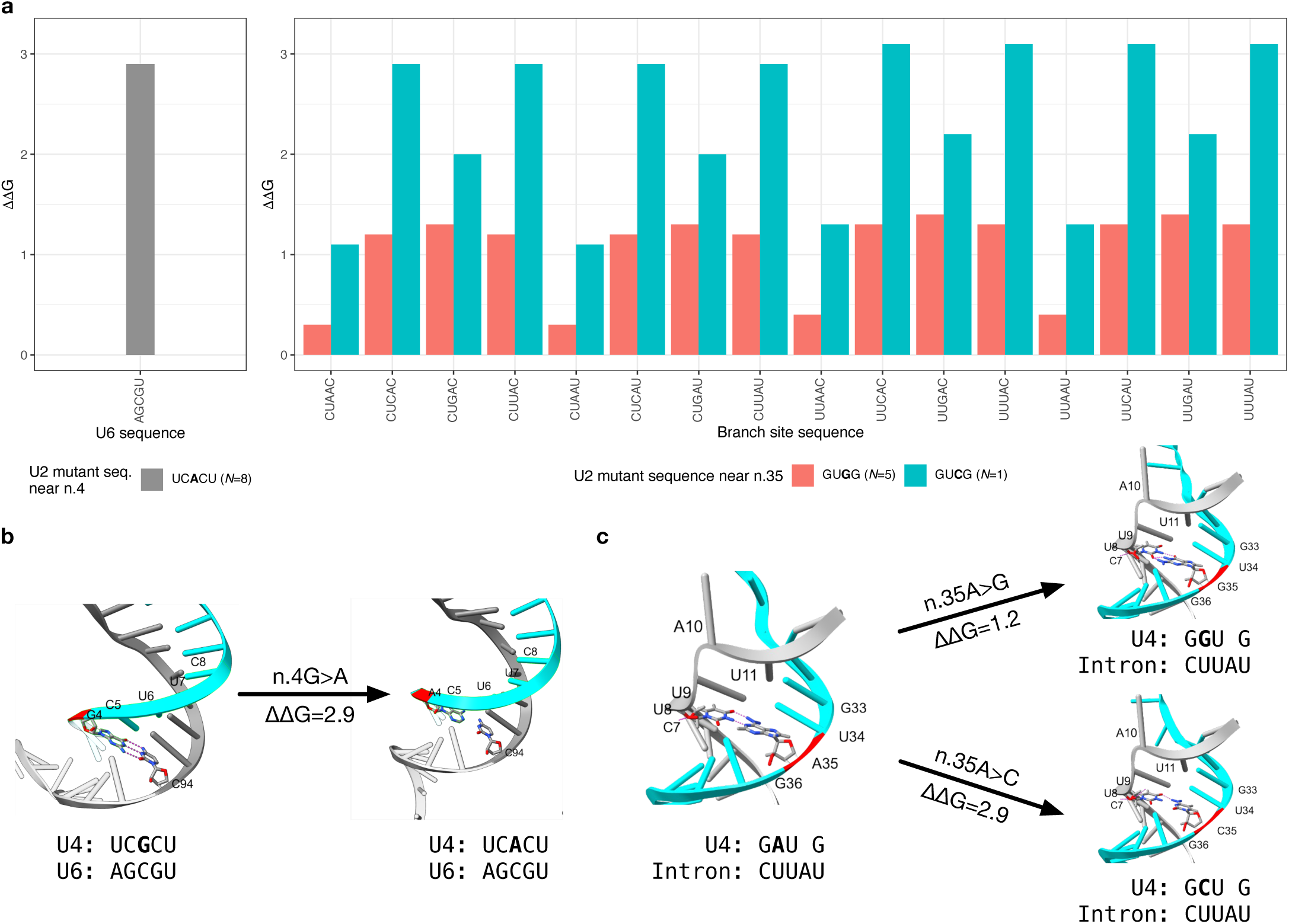
Predicted effects of the mutants on duplex binding stability. **a**, Differential binding stability (ΔΔG) values between U2 and U6 for the A4 mutant allele compared to the reference G4 allele and between U2 and each of 16 branch site sequences consistent with the human YUNAY motif. Each of the substitutions reduces the predicted free energies of association relative to the corresponding reference allele. **b**, For each of the alleles observed at n.4 of *RNU2-2P* (the reference G4 and the mutant A4), a graphical representation of Watson-Crick interactions between the U6-interacting region in U4 (encompassing UCGCU at n.2–6) and the corresponding U6 snRNA region. Hydrogen bonding between cognate nucleotides is depicted with dotted lines. **c**, For each of the germline alleles observed at n.35 (the reference A35 and the mutant G35 and C35 alleles), a graphical representation of Watson-Crick interactions between the branch site recognition region in U4 (GUAG at n.33–36) and an example branch site sequence (CUUAU). Hydrogen bonding between cognate nucleotides is depicted with dotted lines.

**Extended Data Fig. 6.**
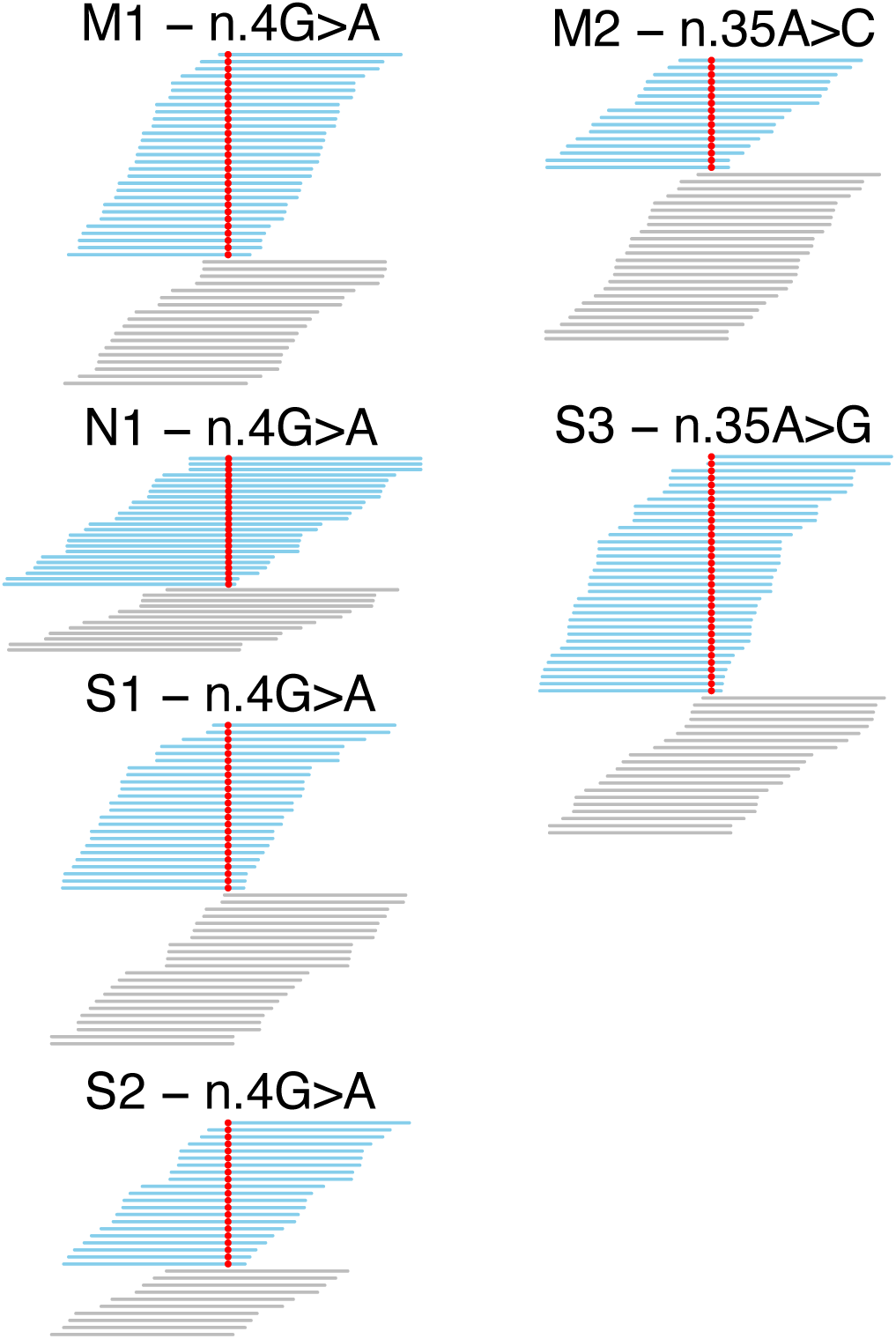
Read pileups in the replication collections. Sequencing read pileups for cases identified in the replication collections. The reads supporting the reference allele are in blue and those supporting the variant allele are in red.

**Extended Data Fig. 7.**
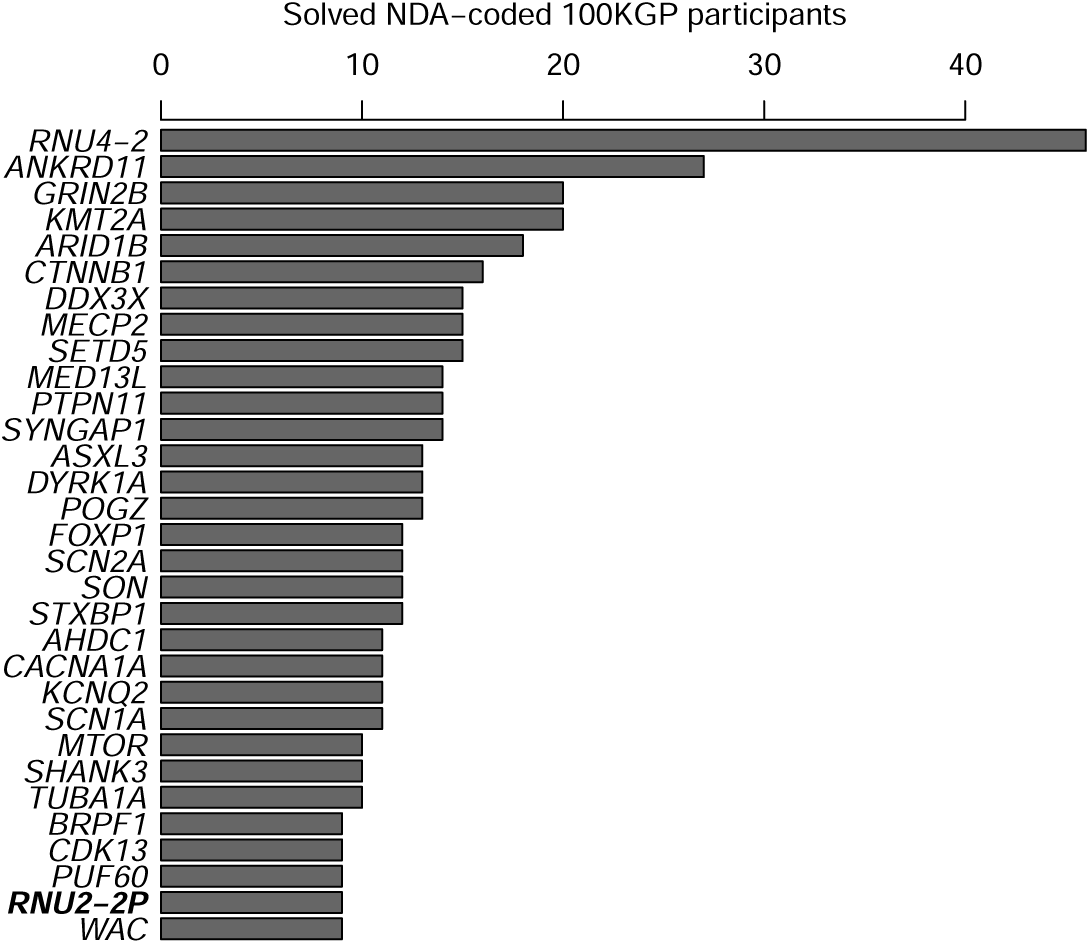
Prevalence in the 100KGP. Out of the 9,112 NDA-coded cases in the 100KGP, the number solved through pathogenic (P) or likely pathogenic (LP) variants in a gene, provided at least 11 cases were diagnosed. In the case of *RNU2-2P*, the number of NDA-coded cases with one of the two recurring de novo variants is shown instead of the number solved with P/LP variants.

**Extended Data Fig. 8.**
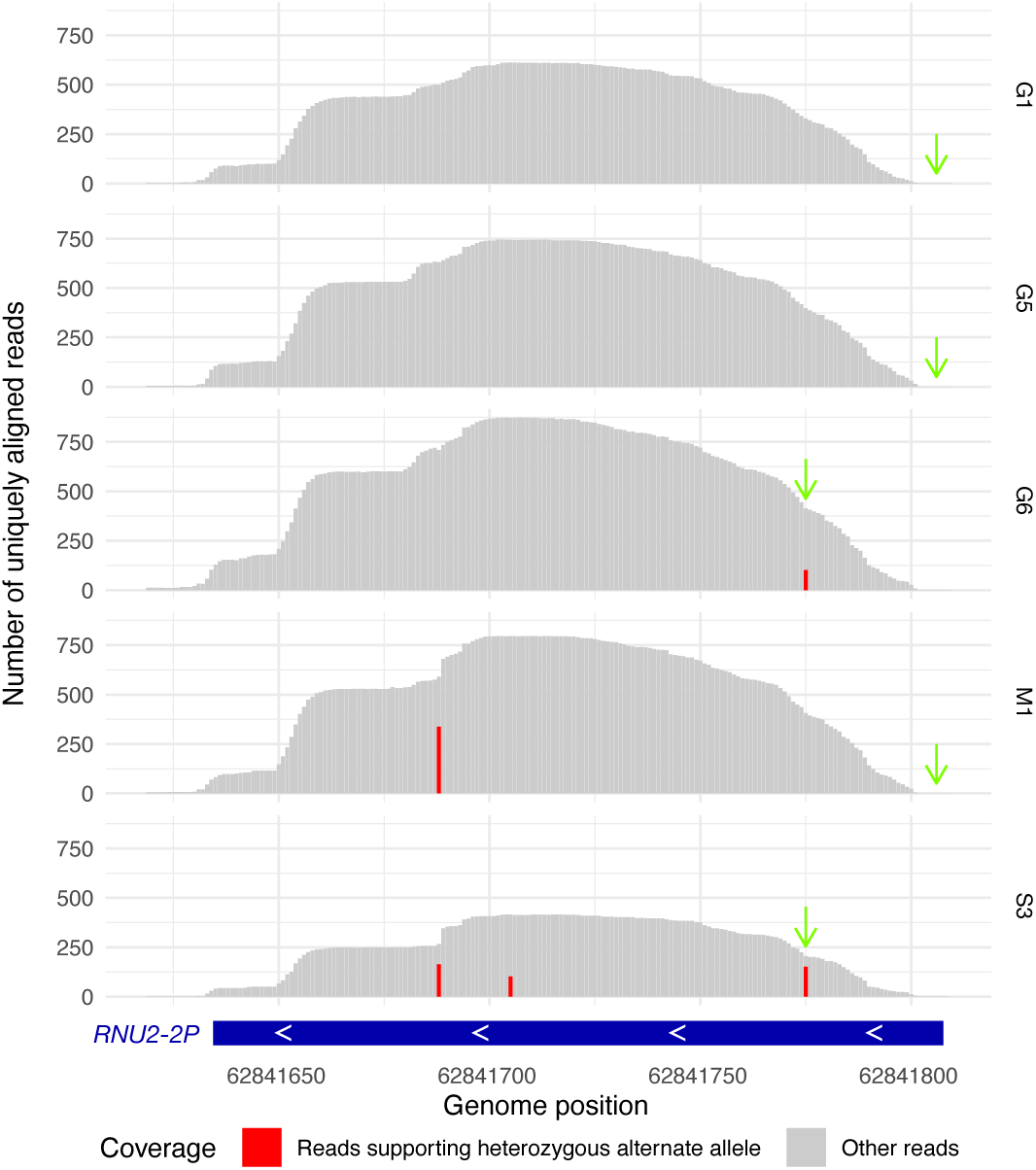
Allele-specific expression of *RNU2-2P* in cases. Coverage of RNA-seq reads from whole blood aligned to the genome near *RNU2-2P* in five cases. The allelic balance at heterozygous sites is shown in red. The locations of the mutant alleles at n.4 and n.35 are indicated with green arrows. The aligned reads overlapping heterozygous sites show that both alleles are expressed robustly in the cases in pedigrees G6, M1 and S3. The cases in pedigrees G1 and G5 were heterozygous only at n.4, where coverage was too low to assess allele-specific expression.

## Supplementary Information

### Case vignettes

`Redacted for medRxiv.`

